# Effectiveness and Safety of Avacopan in Antineutrophil Cytoplasmic Antibody–Associated Vasculitis

**DOI:** 10.64898/2026.07.20.26358270

**Authors:** Lauren Dang, M. Alan Brookhart, Zachary S. Wallace, Alana M. Bozeman, Phuong Pham, Tzu-Chieh Lin, Stephen Motsko, Sam Oh

## Abstract

**Importance:** Avacopan is a small-molecule C5aR1 antagonist used as adjunctive treatment for granulomatosis with polyangiitis (GPA) and microscopic polyangiitis (MPA). Evidence of real-world clinical effectiveness and safety is limited.

**Objective:** Compare effectiveness and describe safety outcomes of avacopan plus SoC versus SoC alone.

**Design:** Retrospective U.S. cohort study with prevalent new-user design emulating sequential nested trials with up to 12-month follow-up. Index dates: first avacopan prescription (avacopan arm); first rituximab or cyclophosphamide claim (SoC arm) during trial window.

**Setting:** Optum Market Clarity administrative claims (October 2015 – June 2025).

**Participants:** Adults receiving rituximab or cyclophosphamide after newly diagnosed or relapsing GPA/MPA. Comparative effectiveness cohort: patients meeting baseline eligibility. Safety cohort: adults with an avacopan prescription, regardless of other eligibility.

**Interventions:** Avacopan plus SoC vs SoC alone.

**Main Outcomes and Measures:** Relapse, prednisone-equivalent daily dose (PEDD) ≤7.5 mg, and serious hepatic event hospitalization were prespecified, whereas cumulative oral glucocorticoid exposure was analyzed post-hoc. A strict hepatic-event screen required [1] acute/subacute hepatic failure, central hemorrhagic liver necrosis, or toxic liver disease and [2] ≥1 code indicative of severe acute liver injury on the same claim; a relaxed hepatic-event screen required either criterion. Standardized mortality ratio and censoring weights accounted for baseline covariates and informative censoring. Treatment effects in the avacopan-treated population were estimated.

**Results:** The effectiveness analysis included 183 avacopan and 4096 SoC index dates. The safety cohort had 828 avacopan users. There were 38 events of relapse in the avacopan arm and 956 in the SoC arm (weighted hazard ratio [95% CI]: 0.81 [0.59, 1.13]). PEDD ≤7.5 mg was numerically more common with avacopan plus SoC at most timepoints. Cumulative glucocorticoid exposure was lower with avacopan plus SoC; between-arm differences exceeded 500 mg from months 5 through 12. No avacopan-exposed patients met the strict hepatic event screen definition. Relaxed hepatic event screen events occurred in 2 (1.1%), 5 (0.6%), and 10 (0.5%) patients in the avacopan effectiveness, avacopan safety, and SoC cohorts, respectively.

**Conclusions and Relevance:** Early evidence from claims data suggests avacopan plus SoC may reduce relapse risk and enable faster glucocorticoid tapering vs SoC alone. Serious hepatic events appear rare.

**Trial Registration:** https://catalogues.ema.europa.eu/node/4889/administrative-details N/A

## INTRODUCTION

Granulomatosis with polyangiitis (GPA) and microscopic polyangiitis (MPA) are 2 types of antineutrophil cytoplasmic antibody (ANCA)–associated vasculitis (AAV), a rare disease often involving life- or organ-threatening manifestations.^1–4^ Treatment for GPA/MPA involves induction with immunosuppressive agents, such as rituximab or cyclophosphamide, in combination with glucocorticoids, followed by prolonged immunosuppressive maintenance therapy.^5–7^

Avacopan is an orally administered, small-molecule C5aR1 antagonist approved for use by the US Food and Drugs Administration as an adjunctive treatment for GPA and MPA in combination with standard therapy, including glucocorticoids.8,9 Avacopan is included in several international guidelines for the treatment of GPA or MPA as part of a strategy to reduce exposure to glucocorticoids.6,7,10 Avacopan was evaluated in the phase 3 ADVOCATE trial (N=331), which assessed remission at week 26 and sustained remission at week 52 among patients receiving induction therapy with rituximab or cyclophosphamide.11 However, clinical trial protocols and populations may not reflect post-approval treatment patterns, and the relatively limited size of trial populations underscores the value of clinical practice evidence from larger and more heterogeneous populations to further inform the benefit-risk profile of avacopan, including relapse, glucocorticoid exposure, and safety.

Post-approval observational studies of avacopan use in patients with GPA and MPA have reported remission, sustained remission, reduced relapse, favorable kidney outcomes,^12–13^ and less cumulative glucocorticoid exposure than traditional regimens.^14–16^ However, much of the available real-world evidence comes from single-arm studies, case series, or small cohorts with limited comparator data, restricting interpretation of the effectiveness of avacopan relative to standard of care (SoC). Real-world studies have also provided safety data.^17–21^ Avacopan is associated with a known risk for hepatotoxicity, which requires monitoring; some cases of vanishing bile duct syndrome, a rare and severe liver injury, have been reported in the post-marketing setting, mostly from Japan.9,22,23

The use of avacopan as adjunctive therapy for GPA/MPA in routine clinical practice provides an opportunity to better characterize its real-world effectiveness and safety relative to SoC alone. This is particularly important because treatment decisions require balancing relapse prevention and glucocorticoid reduction against potential safety risks, including hepatotoxicity. Therefore, this study evaluated the effectiveness and safety of avacopan as an add-on to SoC vs SoC alone among adults with GPA or MPA in a large U.S. real-world cohort.

## METHODS

### Study Design

This retrospective observational comparative effectiveness study was conducted using Optum’s de-identified Market Clarity Data (Optum^®^ Market Clarity), which deterministically links medical and pharmacy claims with electronic health record data, with available data from October 2015 through June 2025. This fit-for-purpose database was selected based on feasibility analyses confirming capture of the claims-based information needed to define treatment exposure, baseline patient characteristics, effectiveness and safety outcomes, and follow-up among patients with AAV. The study was conducted according to a prespecified protocol registered in the European Network of Centres for Pharmacoepidemiology and Pharmacovigilance (ENCePP) electronic register (EUPAS1000000985). Additional details on the manuscript-specific Optum Market Clarity analysis, including cohort construction, treatment indexing, outcome definitions, covariate assessment, weighting, censoring, and clean-room procedures, are provided in the **Supplementary Methods**. The patient inclusion period (ie, indexing period) spanned from October 2021, (corresponding to initial regulatory approval of avacopan in the United States) through March 2025 (3 months before the last date of data availability). A prevalent new-user design that explicitly emulated a sequence of nested trials was used to mitigate confounding and time-alignment biases (**eTable 1**).24,25 This design can generate valid estimates of treatment effects in the situation where the exposure of interest is administered as add-on therapy because it aligns the start of follow-up across treatment strategies, reduces the risk of immortal time bias from requiring patients to remain eligible long enough to initiate avacopan, and avoids comparing patients who may be at different stages of their disease. Together, these design features allowed avacopan initiators to be compared with patients receiving SoC who were eligible to initiate avacopan at the same point in the disease episode.

Sequential nested trials were formed across 14-day intervals starting from each patient’s disease date (newly diagnosed or relapsing GPA/MPA) (**eFigure 1**). Within each interval, avacopan plus SoC arm index dates were defined as the date of the first-ever avacopan prescription. Index dates in the SoC arm were assigned as the first rituximab or cyclophosphamide claim observed during the interval.

Patients could contribute to every trial for which they met eligibility criteria based on information available on or before their trial-specific index date. This approach aims to improve the precision of effect estimates and to emulate trial enrollment procedures that would be based only on eligibility information available before the beginning of follow-up.25 Thus, patients could contribute more than 1 eligible SoC index date and additionally could contribute a later index date to the avacopan + SoC arm. Patients were followed for up to 12 months after the index date.

### Eligibility Criteria

Included in this study were adults aged _≥_18 years, with evidence of a newly diagnosed or relapsing GPA or MPA episode within 125 days prior to index and recent use of rituximab and/or cyclophosphamide (rituximab claim within 90 days and/or cyclophosphamide claim within 45 days) prior to or on the index date. Patients must also have had continuous medical and pharmacy enrollment for ≥12 months prior to index (allowing ≤30-day enrollment gap) and at least 1 day of follow-up. Patients were excluded if they had prior avacopan use before the index date, prior diagnosis of eosinophilic granulomatosis with polyangiitis (EGPA), or prior use of mepolizumab or benralizumab.

### Outcome and Covariate Assessments

Primary endpoints included time to relapse and the proportion of patients reaching a 30-day average oral prednisone-equivalent daily dose (PEDD) of ≤7.5 mg at prespecified timepoints. Incidence of relapse and severe hepatic events were assessed as secondary endpoints. Additional post hoc analyses included monthly 30-day mean PEDD and cumulative glucocorticoid use.

Relapse was assessed using a validated claims-based algorithm that demonstrated 99.2% specificity and 97.7% negative predictive value against expert electronic health record review.26 Relapse was defined as a prednisone claim of ≥20 mg/day for >14 days met by a single claim (not multiple fills combined) within 30 days of AAV diagnosis or relapse manifestation.26 Manifestations included alveolar hemorrhage, respiratory failure, kidney failure, scleritis, retinal exudates/hemorrhage, gangrene, sensorineural deafness, mesenteric ischemia, meningitis, cord lesion, stroke, cranial nerve palsy, sensory neuropathy, and mononeuritis multiplex. To define distinct relapse events, the next eligible relapse must have occurred at least 45 days after the most recent qualifying relapse-related claim date. To reduce false positives from new-onset disease, the first possible relapse must have occurred at least 45 days after the medication associated with new AAV diagnosis and at least 90 days after a GPA/MPA/AAV code. Glucocorticoid exposure was derived from pharmacy claims for oral glucocorticoids by converting doses to PEDD using prespecified conversion factors and day-level exposure construction rules (**Supplementary Methods**). Cumulative glucocorticoid use was defined as the sum of PEDD during follow-up.

The protocol-specified hepatotoxicity/drug-induced liver injury (DILI) endpoint was evaluated as severe hepatic events using a strict screen adapted from a validated claims-based algorithm for severe acute liver injury. The original algorithm was based on principal hospital diagnosis codes, which demonstrated 100% positive predictive value against hepatologist-adjudicated medical records.27 For the current study, the strict hepatic event screen required an inpatient claim with both (1) acute/subacute hepatic failure, central hemorrhagic liver necrosis, or toxic liver disease and (2) ≥1 diagnosis indicative of severe acute liver injury, with diagnoses assessed in any position on the same claim. To evaluate whether additional potentially relevant hospitalized hepatic events may have been missed by the strict screen, a relaxed hepatic event screen was also applied, defined as events meeting either criterion.

Baseline covariates were measured during the 12-month period prior to and inclusive of the index date and included age; sex; time since new AAV diagnosis or relapse; any hospitalization for AAV between a patient’s new or relapsing GPA/MPA diagnosis through their index date; recent glucocorticoid exposure, including pulse glucocorticoids (defined as >125 mg intravenous methylprednisolone summed over 1 day) and high-dose oral glucocorticoids (defined as daily prednisone equivalent >20 mg); specific AAV manifestations (ie, diffuse alveolar hemorrhage, chronic kidney disease, acute kidney injury, dialysis, or kidney transplant); and recent use of rituximab/cyclophosphamide.

### Statistical Analysis

Logistic regression was used to estimate the propensity score, defined as the probability of initiating avacopan (avacopan plus SoC) vs remaining on SoC alone (SoC) on the index date, conditional on time from the new or relapsing disease date (modeled using a cubic spline) and other baseline covariates, pooled across all 14-day trials. To control for confounding, standardized mortality ratio (SMR) weights were applied to reweight the SoC arm to the covariate distribution of avacopan initiators, thereby providing estimates of effects in the avacopan-treated population. Covariate balance between treatment groups was assessed before and after SMR weighting using standardized mean differences (SMDs), with SMDs <0.1 considered indicative of adequate balance; additional diagnostics included propensity score overlap and weight distributions. Inverse probability of censoring (IPC) weights were used to account for potentially informative censoring. To assess residual bias, negative control outcomes were evaluated after propensity score and IPC weighting and included screening mammography (women) or prostate cancer screening (men), and wellness visits.28,29 Trials were pooled to estimate overall effects as SMR and IPC–weighted hazard ratios, risk ratios, or differences in mean values of continuous endpoints. As patients could contribute more than 1 eligible index treatment date, confidence intervals were constructed using a grouped patient-level bootstrap to account for within-patient correlation.30

Primary effectiveness analyses followed a modified intent-to-treat framework, in which patients receiving SoC were censored if they filled an avacopan prescription within 30 days after index. This approach is intended to account for short delays between treatment intent and the observed avacopan prescription, particularly for patients whose treatment may have been intended to start at the index date but was recorded shortly thereafter. Patients receiving SoC or avacopan plus SoC who met all eligibility criteria formed the effectiveness cohort. For each safety endpoint in the effectiveness cohort, the cumulative incidence within 12 months following the index date was estimated separately by arm using baseline propensity score SMR weights without IPC weights. An expanded safety cohort included all adults in the closed-claims data with a recorded avacopan prescription during observed enrollment within the study period. For this expanded safety cohort, the number and proportion of patients with a safety event within 12 months following the first recorded avacopan prescription were summarized descriptively.

### Clean-Room Process

The study was designed and executed using a prespecified and staged analytic framework and strict clean-room procedures.31,32 These procedures included predefined analytic checkpoints, locking of the final statistical analysis plan before primary and secondary outcome analyses, restricted access to analytic data, masking of treatment-group information where appropriate, and maintenance of an audit trail and decision log. This approach was intended to ensure that design refinements and analytic decisions were made without knowledge of their impact on treatment effect estimates.

## RESULTS

The avacopan plus SoC effectiveness cohort included 183 patients, each contributing 1 index treatment date. The SoC cohort included 2103 unique patients contributing 4096 eligible index treatment dates based on rituximab or cyclophosphamide claims (**eFigure 2**). The expanded avacopan safety cohort included 828 adult patients with a recorded avacopan prescription. Before weighting, the avacopan + SoC cohort had greater baseline disease severity and glucocorticoid exposure than the SoC cohort (**Table 1**), as reflected by higher recent PEDD, more high-dose oral glucocorticoid use, and more frequent pulmonary and kidney manifestations. Baseline characteristics were balanced after weighting, with all standardized mean differences <0.1 (**Table 1, eFigure 3**).

**Table 1.** Demographics and Baseline Characteristics.

| Characteristic | Unweighted |  |  | Weighted |  |  |
| --- | --- | --- | --- | --- | --- | --- |
|  | SoC<br>(n=4096) | Avacopan Plus<br>SoC<br>(n=183) | SMD | SoC<br>(n=4279<sup>a</sup>) | Avacopan Plus<br>SoC<br>(n=4279<sup>a</sup>) | SMD |
| **Demographics and disease characteristics** |  |  |  |  |  |  |
| Age, mean (SD), years | 58.9 (15.7) | 54.2 (16.2) | 0.299 | 54.2 (16.0) | 54.2 (16.1) | 0.002 |
| Female, No. (%) | 2306 (56.3) | 115 (62.8) | 0.134 | 2682 (62.7) | 2689 (62.8) | 0.003 |
| Vasculitis disease status,<br>No. (%) |  |  | 0.037 |  |  | 0.018 |
| Newly diagnosed | 2912 (71.1) | 127 (69.4) |  | 2935 (68.6) | 2970 (69.4) |  |
| Relapsing | 1184 (28.9) | 56 (30.6) |  | 1344 (31.4) | 1309 (30.6) |  |
| Days from disease to<br>index, mean (SD) | 45.2 (31.5) | 55.4 (27.7) | 0.345 | 55.1 (27.7) | 55.4 (27.6) | −0.009 |
| **Comorbidities** |  |  |  |  |  |  |
| Charlson Comorbidity<br>Index, mean (SD) | 2.8 (2.2) | 2.6 (2.0) | 0.079 | 2.6 (2.2) | 2.6 (2.0) | −0.011 |
| Type 2 diabetes, No. (%) | 864 (21.1) | 29 (15.9) | 0.136 | 678 (15.8) | 678 (15.8) | 0.0003 |
| Hypertension, No. (%) | 2578 (62.9) | 101 (55.2) | 0.158 | 2338 (54.6) | 2362 (55.2) | 0.011 |
| **Manifestations** |  |  |  |  |  |  |
| Diffuse alveolar<br>hemorrhage, No. (%) | 328 (8.0) | 23 (12.6) | 0.151 | 544 (12.7) | 538 (12.6) | 0.005 |
| Acute kidney injury,<br>No. (%) | 1837 (44.9) | 95 (51.9) | 0.142 | 2164 (50.6) | 2221 (51.9) | 0.027 |
| Chronic kidney disease,<br>No. (%) | 1856 (45.3) | 78 (42.6) | 0.054 | 1794 (41.9) | 1824 (42.6) | 0.014 |
| Other kidney diseases,<br>No. (%) | 1965 (48.0) | 103 (56.3) | 0.167 | 2391 (55.9) | 2408 (56.3) | 0.008 |
|  | SoC<br>(n=4096) | Avacopan Plus<br>SoC<br>(n=183) | SMD | SoC<br>(n=4279<sup>a</sup>) | Avacopan Plus<br>SoC<br>(n=4279<sup>a</sup>) | SMD |
| **Recent glucocorticoid exposures<sup>b</sup>** |  |  |  |  |  |  |
| Average PEDD, mean (SD), mg | 19.4 (18.2) | 29.3 (21.6) | 0.499 | 29.2 (21.9) | 29.3 (21.5) | -0.007 |
| Average PEDD ≤7.5 mg, No. (%) | 1246 (30.4) | 35 (19.1) | 0.264 | 750 (17.5) | 818 (19.1) | 0.037 |
| Pulse-dose glucocorticoids, No. (%) | 406 (9.9) | 24 (13.1) | 0.101 | 582 (13.6) | 561 (13.1) | 0.015 |
| High-dose oral glucocorticoids, No. (%) | 3112 (76.0) | 159 (86.9) | 0.283 | 3701 (86.5) | 3718 (86.9) | 0.010 |
| **Background therapy** |  |  |  |  |  |  |
| Rituximab<sup>c</sup> | 3664 (89.5) | 173 (94.5) | 0.188 | 4043 (94.5) | 4045 (94.5) | 0.002 |
| Cyclophosphamide<sup>c</sup> | 546 (13.3) | 20 (10.9) | 0.074 | 487 (11.4) | 468 (10.9) | 0.014 |
| Other DMARDs<sup>d</sup> | 526 (12.8) | 18 (9.8) | 0.095 | 417 (9.8) | 421 (9.8) | 0.003 |
| **Healthcare resource utilization** |  |  |  |  |  |  |
| Hospitalization for AAV, No. (%) | 1319 (32.2) | 87 (47.5) | 0.317 | 2031 (47.5) | 2034 (47.5) | 0.002 |
| Outpatient visits, mean (SD) | 17.6 (11.9) | 17.5 (10.6) | 0.004 | 17.5 (10.9) | 17.5 (10.6) | -0.0002 |
| Inpatient/ER visits, mean (SD) | 2.3 (4.1) | 2.5 (3.0) | 0.044 | 2.5 (4.1) | 2.5 (3.0) | 0.013 |

<sup>a</sup>Weighted n is equal to the sum of the normalized standardized mortality ratio weights, which are normalized to sum to the total sample size.
<sup>b</sup>Recent exposures assessed in the 60 days prior to and inclusive of the index date.
<sup>c</sup>Exposures to rituximab and cyclophosphamide were assessed in the 90 and 45 days prior to and inclusive of the index date, respectively.
<sup>d</sup>Any claim for methotrexate, azathioprine, mycophenolic acid, and/or mycophenolate mofetil.
AAV, antineutrophil cytoplasmic antibody-associated vasculitis; DMARD, disease-modifying antirheumatic drug; ER, emergency room; PEDD, prednisone-equivalent daily dose; SD, standard deviation; SMD, standardized mean difference; SoC, standard of care.

There were 38 relapses in the avacopan plus SoC arm and 956 in the SoC arm (unweighted cumulative 12-month risk: 23.9 vs 28.0, 12-month risk ratio [RR] [95% CI] = 0.85 [0.62, 1.19]; weighted cumulative 12-month risk: 24.3 vs 28.1, 12-month RR [95% CI] = 0.87 [0.66, 1.27]) (**Figure 1**). Unweighted and weighted hazard ratios (95% CI) for first relapse were 0.86 (0.63, 1.25) and 0.81 (0.59, 1.13), respectively. In both the unweighted and weighted analyses, cumulative relapse risk was similar early after index, with the curves beginning to diverge after approximately 2 to 3 months; lower cumulative risk in the avacopan plus SoC arm was observed by approximately day 90 and persisted through 12 months.

**Figure 1.**
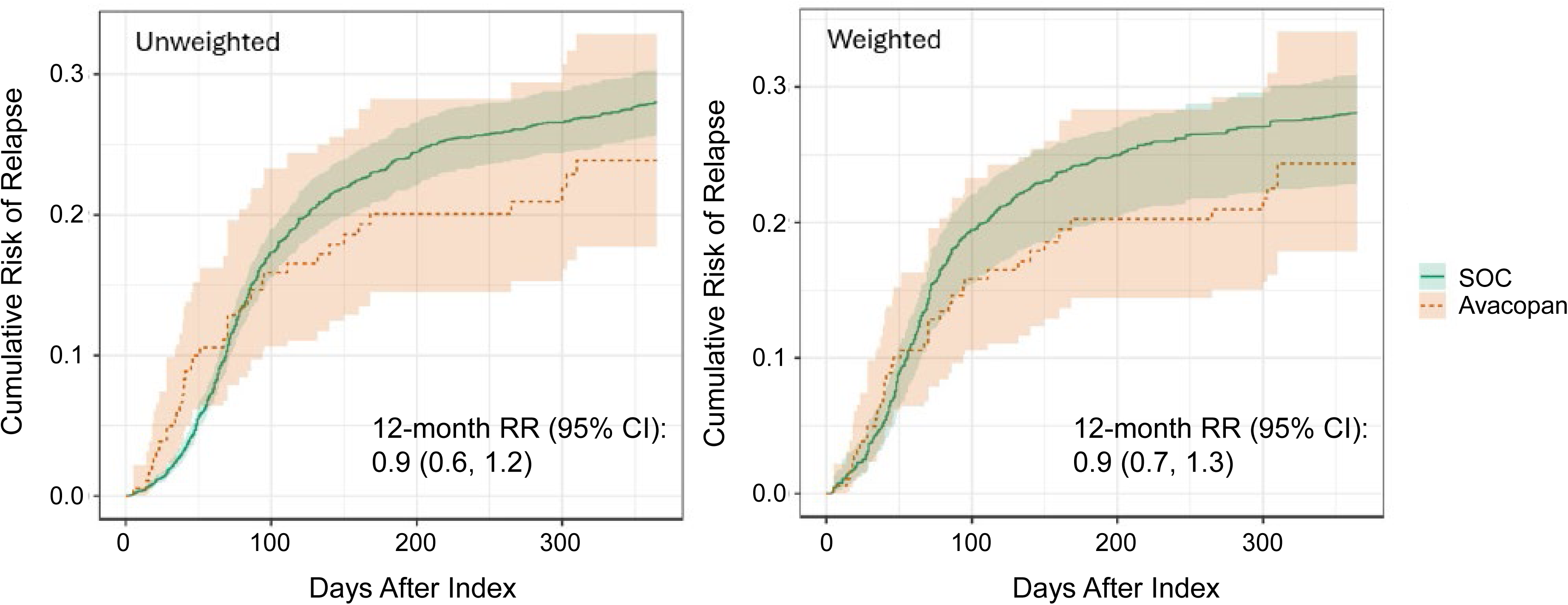
Cumulative Risk of Relapse by 12 Months. Unweighted (left panel) and weighted (right panel) cumulative risk of relapse by 12 months between avacopan plus SoC and SoC arms. RR, relative risk; SoC, standard of care.

In the unweighted analysis, avacopan plus SoC was associated with a numerically higher estimated probability of achieving 30-day average PEDD ≤7.5 mg vs SoC through Day 270, with similar estimates at Day 365. In the weighted analysis, estimates numerically favored avacopan plus SoC at each prespecified timepoint (**Table 2**). A post-hoc analysis of monthly 30-day mean PEDD showed reductions in both arms over time, with greater reductions in the avacopan plus SoC arm than the SoC arm occurring during the first 2 to 5 months after index and with convergence later in follow-up as glucocorticoid exposure declined in both arms (**Figure 2**). Mean cumulative glucocorticoid exposure was lower in the avacopan vs SoC arm over follow-up, with between-arm differences increasing over time. A weighted difference >500 mg in cumulative glucocorticoid exposure was achieved by month 5 and maintained through month 12 (**Figure 3**). The unweighted and weighted mean differences (95% CI) at 6 months were −399.1 mg (−701.6, −27.8) and −559.7 mg (−814.2, −1.9), respectively; at 12 months, the unweighted and weighted mean differences (95% CI) were −517.0 mg (−944.3, −54.2) and −633.3 mg (−984.7, 17.7), respectively.

**Figure 2.**
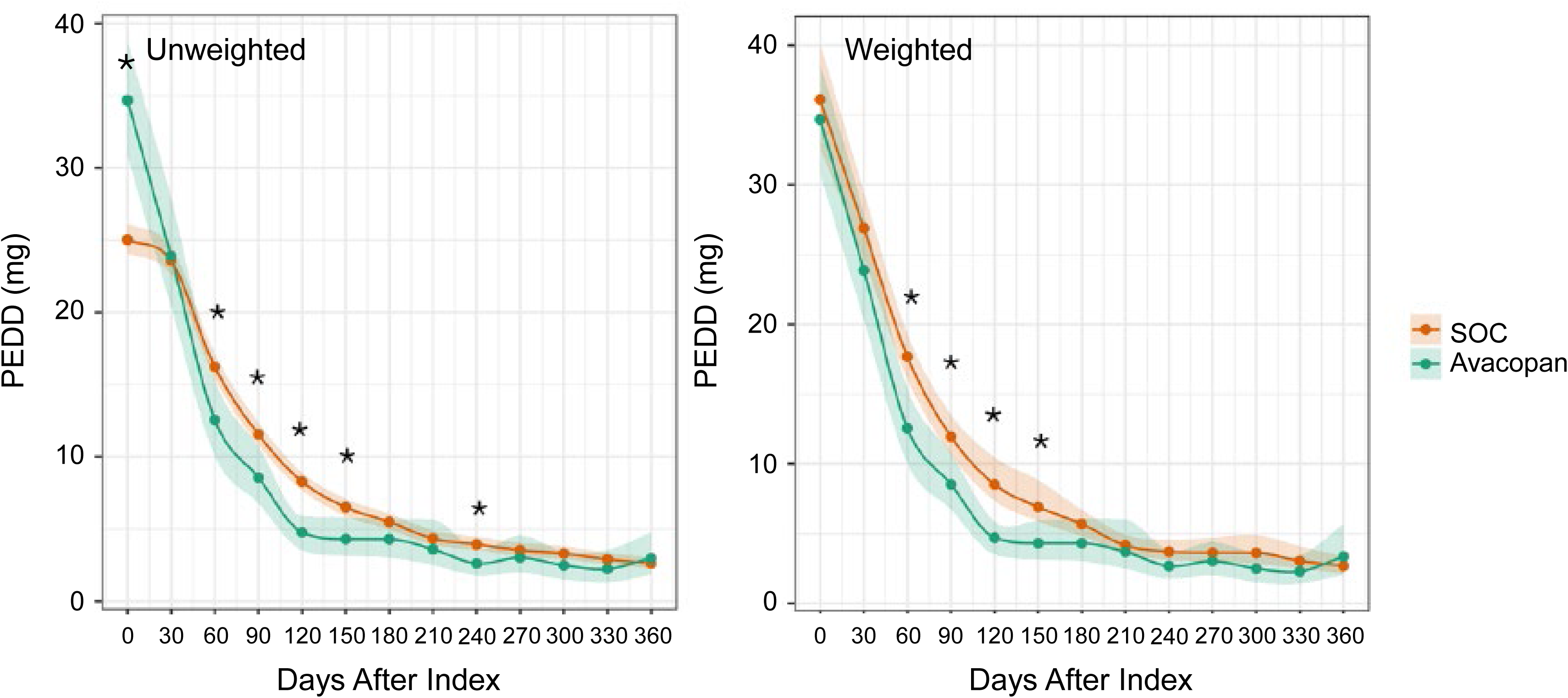
Mean 30-day Average PEDD at Monthly Intervals. \**P*<.05 for difference in PEDD between arms. Unweighted (left panel) and weighted (right panel) mean 30-day average PEDD. Shading indicates 95% CI. CI, confidence interval; PEDD, prednisone-equivalent daily dose; SoC, standard of care

**Figure 3.**
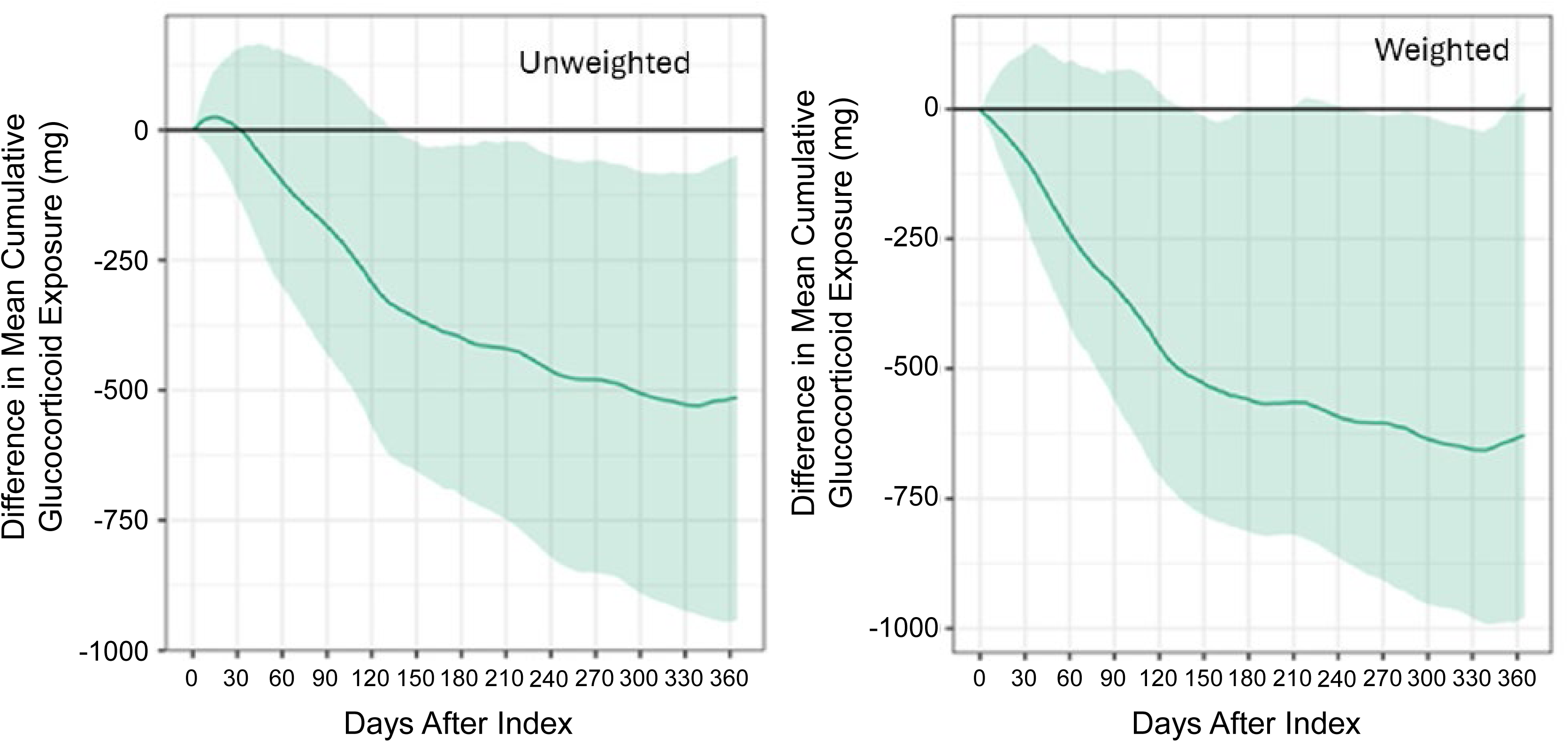
Cumulative Glucocorticoid Exposure Difference Between Avacopan plus SoC and SoC Arms. Unweighted (left panel) and weighted (right panel) difference in mean cumulative oral glucocorticoid exposure. SoC, standard of care

**Table 2.** Probability Ratio for Achieving 30-Day Average PEDD ≤7.5 mg.

|  | Unweighted |  |  | Weighted |  |  |
| --- | --- | --- | --- | --- | --- | --- |
| Days | SoC<br>Percent<br>(95% CI) | Avacopan Plus<br>SoC<br>Percent<br>(95% CI) | PR<sup>a</sup><br>(95% CI) | SoC<br>Percent<br>(95% CI) | Avacopan Plus<br>SoC<br>Percent<br>(95% CI) | PR<sup>a</sup><br>(95% CI) |
| 90 | 58.38<br>(56.21, 60.82) | 63.03<br>(52.15, 75.21) | 1.08<br>(0.90, 1.32) | 57.25<br>(48.60, 60.48) | 62.96<br>(52.31, 74.80) | 1.10<br>(0.94, 1.43) |
| 120 | 67.29<br>(64.74, 69.74) | 77.71<br>(66.32, 91.87) | 1.15<br>(0.98, 1.38) | 67.74<br>(58.24, 71.69) | 77.67<br>(66.80, 91.66) | 1.15<br>(1.00, 1.51) |
| 180 | 78.01<br>(75.61, 79.97) | 79.10<br>(66.66, 92.25) | 1.01<br>(0.85, 1.21) | 74.79<br>(63.89, 78.64) | 78.52<br>(66.59, 91.94) | 1.05<br>(0.91, 1.37) |
| 270 | 85.77<br>(83.50, 87.54) | 90.43<br>(78.65, 100.00) | 1.05<br>(0.91, 1.26) | 80.88<br>(69.40, 84.86) | 91.02<br>(78.46, 100.00) | 1.13<br>(0.98, 1.45) |
| 365 | 88.98<br>(86.828, 90.70) | 86.81<br>(74.09, 100.00) | 0.98<br>(0.83, 1.17) | 81.11<br>(69.38, 87.98) | 83.75<br>(71.08, 99.61) | 1.03<br>(0.86, 1.31) |

<sup>a</sup>Probability ratios compare avacopan plus SoC vs SoC. A probability ratio > 1 indicates a higher probability of having 30-day average PEDD $\leq 7.5$ mg.
CI, confidence interval; PEDD, prednisone-equivalent daily dose; PR, probability ratio; SoC, standard of care.

Using the protocol-specified strict hepatic event screen, no serious hepatic events were identified in the avacopan effectiveness or safety cohorts, and 2 events (0.1%) were identified in the SoC cohort. Under the relaxed hepatic event screen, 2 (1.1%) patients were identified in the avacopan plus SoC effectiveness cohort, 5 (0.6%) in the avacopan safety cohort, and 10 (0.5%) in the SoC cohort.

## DISCUSSION

This was among the first comparative effectiveness studies of avacopan plus SoC versus SoC alone conducted in the current treatment era. In this study, patients prescribed avacopan as add-on to SoC had a numerically lower relative hazard of relapse after weighting, achieved PEDD ≤ 7.5mg/d faster, and experienced lower glucocorticoid exposure than SoC alone. Although the relapse estimate did not reach statistical significance, the direction of effect is clinically relevant because relapse contributes to organ damage due to disease activity, and the lower estimated relapse risk occurred despite lower glucocorticoid exposure in the avacopan plus SoC arm.

Reduction in glucocorticoid exposure is a key treatment goal in clinical practice in order to reduce glucocorticoid-related toxicity.6,7,10 Serious hepatic events were rare in the avacopan plus SoC arm, the expanded avacopan safety cohort, and the SoC arm.

Prior real-world studies have generally reported favorable remission, relapse, kidney, and glucocorticoid-sparing outcomes among patients receiving avacopan, but much of the available evidence has been limited by small comparative cohorts, historical-control analyses, and single-arm studies.16,^33–35^ This study extends prior evidence by evaluating avacopan plus SoC versus contemporary SoC alone using a prevalent new-user design with sequential nested trial emulations. The direction of the findings is also generally consistent with observations from the phase 3 ADVOCATE trial, in which avacopan was evaluated with standard immunosuppressive therapy and found to be associated with a reduction in the risk of relapse despite an 81% reduction in median glucocorticoid exposure when compared to a prednisone taper.36 The smaller magnitude of relapse reduction observed in this study compared with ADVOCATE may reflect differences between protocolized clinical trial care and routine practice, including evolving SoC treatment patterns, use of maintenance rituximab, non-protocolized glucocorticoid tapering, and post-index changes in exposure that were not captured under the modified intent-to-treat framework.

The glucocorticoid outcomes should be interpreted in the context of differences between protocolized clinical trial glucocorticoid tapering and clinician-directed glucocorticoid management in routine practice. Real-world studies by Patel et al. recently illustrated substantial variability after avacopan initiation, including ongoing glucocorticoid use in some patients, while also suggesting that shorter tapers can reduce glucocorticoid exposure with similar disease outcomes compared with longer tapers.37,38 In the present study, avacopan + SoC was associated with lower cumulative glucocorticoid exposure than SoC alone despite this variability.

Hepatotoxicity is a known risk associated with avacopan treatment and is included in product labeling.9 In a meta-analysis of 16 real-world studies, the pooled proportion of patients with hepatotoxicity events was 0.06 (95% CI, 0.03–0.14), although the authors noted substantial heterogeneity, with higher proportions predominantly observed in Japanese cohorts.12 The US Food and Drug Administration issued a Drug Safety Communication regarding serious liver injury cases with avacopan, including some cases of DILI involving vanishing bile duct syndrome in the postmarketing setting. Importantly, the majority of the observed DILI cases were reported from Japan.39 In this large US-based cohort, the overall incidence of serious hepatic events was very low in the avacopan plus SoC arm and in the expanded avacopan safety cohort using both a strict and more relaxed screen for hospitalized hepatic events. The low incidence of serious hepatic events in the SoC arm suggests a low background rate of hospitalized hepatic events among patients with AAV initially treated with SoC alone. Similarly, a separate real-world analysis of patients with AAV found similar DILI incidence after propensity score matching among patients treated with versus without avacopan (2.0% vs 1.9%).40 The relaxed hepatic event screen identified a small number of hospitalized hepatic events among avacopan-exposed patients, and these events occurred at a frequency similar to that observed in the SoC cohort.

### Limitations

This analysis has some limitations. As a claims-based analysis, the study may have been subject to variability in coding practices and incomplete capture of clinical information, including potential undercoding of some diagnoses, disease manifestations, and outcomes. Important clinical measures of disease activity, including the Birmingham Vasculitis Activity Score (BVAS), are not available in claims data. Accordingly, active disease burden at baseline could not be assessed directly, and disease severity had to be inferred from proxy measures captured in claims. Because the primary effectiveness analyses used a modified intent-to-treat framework, post-index treatment changes after the initial treatment-strategy window, including avacopan discontinuation, were not explicitly accounted for; therefore, the observed effectiveness estimates are expected to be conservative. However, this approach allowed all safety events during the 12-month follow-up to be captured. Finally, although this study used a large real-world database, the avacopan effectiveness cohort was modest in size, limiting precision for some outcomes. Additional analyses with larger avacopan-treated populations are needed to confirm these findings.

### Strengths

This study has several strengths relevant to interpretation. The analysis used a large longitudinal administrative claims database that captures medical encounters, pharmacy dispensing, treatments, and outcomes among patients receiving care in routine clinical practice. As described in the Methods, feasibility analyses demonstrated that the Optum data were fit-for-use for this specific study question since data on exposures, baseline characteristics, clinical outcomes, and safety events were reliably captured and that patients with AAV could be validly identified. The study adhered to pharmacoepidemiologic best practices and ICH M14 guidance.41,42 The study was governed by a protocol that was prespecified and registered on the ENCePP electronic register; followed the target trial emulation framework with contemporary causal inference methods; and used staged analytic procedures and clean-room safeguards to reduce the risk of bias from data-driven decision-making. Covariate balance in this analysis improved substantially after weighting, supporting the comparability of the adjusted populations. The use of negative control outcomes provided an additional assessment of potential residual bias, including bias from unmeasured or incompletely measured confounding. A validated algorithm was used for relapse, and hepatotoxicity/DILI was operationalized by adapting a previously validated claims-based algorithm.26,27

## CONCLUSION

Patients with GPA or MPA receiving avacopan as an add-on to SoC in clinical practice had numerically lower risk of relapses and reduced glucocorticoid exposure than patients receiving SoC alone. Serious hepatic events were rare in both arms. These findings provide additional evidence to support a favorable benefit-risk profile for avacopan.

## Data Availability

The patient-level data underlying this study are not publicly available because they contain proprietary elements owned by Optum and were accessed under license for the current study. Broad disclosure or public sharing is not permitted under the terms of the data license. Any third-party access is subject to Optum's data access requirements, including data security and privacy protocols and a license agreement governing use of the data.

## ACKNOWLEDGMENTS

Writing and editorial support was funded by Amgen Inc. and provided by Rebecca Lane, PhD, of Peloton Advantage, LLC, an OPEN Health company, and funded by Amgen Inc.

## Author Contributions

Lauren Dang and Sam Oh had full access to the data and take full responsibility for the integrity of the data and the accuracy of the data analysis.

Concept and design: All authors

Acquisition, analysis, or interpretation of data: All authors

Drafting of the manuscript: All authors

Critical review of the manuscript for important intellectual content: All authors

Statistical analysis: Lauren Dang; M. Alan Brookhart

Obtained funding: N/A

Administrative, technical, or material support: N/A

Supervision: Lauren Dang; Tzu-Chieh Lin; Stephen Motsko; Sam Oh

Other - Study management committee member: N/A

