## Supplement for "Effectiveness and Safety of Avacopan in Antineutrophil Cytoplasmic Antibody–Associated Vasculitis"

**Supplement 1**

**Supplementary Methods**

**eTable 1. Modified ITT Estimands for Comparative Effectiveness and Safety**

**eFigure 1.** **Study Design**

**eFigure 2.** **Patient Attrition**

**eFigure 3.** **Standardized Mean Differences of Key Baseline Characteristics Among the Avacopan Plus SoC and SoC Arms Before and After Propensity Score Weighting**

**References**

**Supplementary Methods**

**Purpose and Scope**

The full study protocol was registered in the European Network of Centres for Pharmacoepidemiology and Pharmacovigilance (ENCePP) electronic register. This supplementary methods document summarizes the Optum Market Clarity analyses reported in the accompanying manuscript and provides additional methodologic detail for those analyses, without reproducing the full parent protocol or statistical analysis plan. The parent protocol and statistical analysis plan include additional planned analyses, data sources, and endpoints that are not reported in the accompanying manuscript and are outside the scope of this supplement. Those analyses will be addressed in future reports.

**Data Source and Study Period**

The study used Optum Market Clarity, a linked administrative claims and electronic health record database. The analyses used the administrative claims component to capture enrollment, pharmacy dispensing, medical procedures, diagnoses, and outcomes. Available data spanned October 1, 2015, through June 2025. The indexing period began October 1, 2021, corresponding to availability of avacopan in the United States, and ended March 31, 2025, 3 months before the end of available data.

**Study Design and Index Date Assignment**

This is a retrospective observational comparative effectiveness study of avacopan + standard of care (SoC) vs SoC among adults with newly diagnosed or relapsing granulomatosis with polyangiitis (GPA) or microscopic polyangiitis (MPA). A prevalent new-user design with sequential nested trials was used to emulate a target trial framework and to mitigate confounding and time-alignment biases. Details of the target trial emulation are provided in Supplementary Table 1.

Sequential trials were emulated every 14 days from each patient's disease date through 125 days after the disease date. Within each trial, eligible patients could contribute an index date if they met trial-specific eligibility criteria based on information available on or before that index date.

For the avacopan + SoC arm, the index date was the date of the first avacopan prescription occurring on or after the disease date and before any subsequent disease date. For the SoC arm, the index date was the earliest rituximab or cyclophosphamide claim during the relevant 14-day trial window. Patients could contribute more than one eligible SoC index date and, if avacopan was initiated later, could subsequently contribute an avacopan index date. Follow-up began the day after the index date and continued until the outcome of interest, disenrollment, death, end of administrative follow-up, or 12 months, as applicable. Patients were also censored from the SoC arm at the time of avacopan initiation if they initiated avacopan within 30 days after index, consistent with the treatment strategies of the target trial.

**Cohort Eligibility**

The primary analysis set included adults aged at least 18 years with newly diagnosed or relapsing GPA/MPA within 125 days before index and recent use of rituximab or cyclophosphamide, defined as a rituximab claim within 90 days before or on index or a cyclophosphamide claim within 45 days before or on index. Patients were required to have at least 12 months of continuous medical and pharmacy enrollment before index, allowing gaps of no more than 30 days, and at least 1 day of follow-up after index.

Patients were excluded if they had avacopan use before index, avacopan use on the index date in the SoC arm, evidence of eosinophilic granulomatosis with polyangiitis, use of mepolizumab or benralizumab, a death date before or on the index date, no qualifying new or relapsing GPA/MPA event, or an index date after March 31, 2025. Because no patients prescribed avacopan had unknown gender or insurance in Optum, SoC records with unknown gender or insurance were excluded.

**Disease Episode Definitions**

New GPA/MPA was defined by 1 inpatient claim or 2 outpatient claims, separated by 14 to 365 days, with a diagnosis code for GPA, MPA, or antineutrophil cytoplasmic antibody (ANCA)–associated vasculitis (AAV). The qualifying inpatient diagnosis date, or the first of the 2 qualifying outpatient diagnosis dates, was required to occur within ±120 days of a claim for glucocorticoids or another immunosuppressive medication. The earliest diagnosis date meeting these criteria was considered the new disease date.

Relapsing GPA/MPA was identified using a claims-based algorithm requiring a prednisone claim of at least 20 mg/day for more than 14 days, met by a single claim, with the prednisone start date within ±30 days of a qualifying GPA, MPA, AAV, or major relapse-associated manifestation code. Relapse-associated manifestations included: alveolar hemorrhage, respiratory failure, kidney failure, scleritis, retinal exudates/hemorrhage, gangrene, sensorineural deafness, mesenteric ischemia, meningitis, cord lesion, stroke, cranial nerve palsy, sensory neuropathy, mononeuritis multiplex. To avoid misclassifying new-onset or baseline active disease as relapse, a prior GPA/MPA/AAV code was required at least 90 days before the relapse event date, and patients were not eligible for relapse until at least 45 days after the treatment date associated with their new diagnosis. After one relapse, the next eligible relapse was required to occur at least 45 days after the last qualifying ICD-10 code (paired with a qualifying prednisone code).

**Outcomes**

| Outcome | Classification | Effect Measure |
| --- | --- | --- |
| Time to first relapse | Primary | Hazard ratio for first relapse over 12 months. |
| 30-day average PEDD ≤7.5 mg | Primary | Ratio of proportions with average PEDD ≤7.5 mg during the 30 days before days 90, 120, 180, 270, and 365 after index. |
| Relapse within 12 months | Secondary | Cumulative risk, risk ratio, and risk difference by 12 months. |
| Serious hepatic events | Secondary safety | Cumulative risk of strict and relaxed hospitalized hepatic event screens within 12 months. |
| Monthly 30-day average PEDD | Post hoc | Difference in mean 30-day average PEDD at 30-day intervals from day 30 through day 360. |
| Cumulative glucocorticoid exposure | Post hoc | Difference in mean cumulative prednisone-equivalent exposure at 30-day intervals from day 30 through day 360. |

Oral glucocorticoid exposure was derived from pharmacy claims and converted to prednisone-equivalent daily dose (PEDD). Each claim was converted to a prednisone-equivalent dose using drug-specific conversion factors, divided by days' supply to assign a daily dose, and summed across same-day claims. A non-stockpiling approach was used for oral glucocorticoids, allowing overlaps between medication dispensings.

**Hepatic Event Definitions**

The protocol-specified hepatotoxicity/drug-induced liver injury endpoint was operationalized as serious hepatic events using a strict screen adapted from a validated claims-based algorithm for severe acute liver injury. The strict screen required an inpatient claim with both: (1) acute/subacute hepatic failure, central hemorrhagic necrosis of the liver, or toxic liver disease; and (2) at least one additional diagnosis indicative of severe acute liver injury on the same claim. Diagnoses were assessed in any position on the same inpatient claim. A relaxed screen was also applied, defined as an inpatient claim meeting either of these criteria.

**Baseline Covariates and Propensity Score Variables**

Baseline covariates were assessed during the 365 days before index through the index date, unless otherwise specified. Diagnosis-based covariates generally required 1 inpatient or 2 outpatient claims, and procedure-based covariates generally required 1 inpatient or 1 outpatient claim, with exceptions defined in the analysis specifications. Final action, paid claims were used for all variables.

Propensity score and censoring models included demographic characteristics, calendar time, insurance type, disease timing, healthcare utilization, disease severity proxies, glucocorticoid exposure, organ involvement, comorbidities, prior and recent immunosuppressive medication use, and healthcare access or engagement variables. The final propensity score model used to define the standardized mortality ratio weights included the following variables or variable groups:

- Age, gender, race, index year (as a continuous variable), and insurance type.
- Time from disease date to index, modeled using a natural cubic spline with 3 degrees of freedom.
- AAV-related hospitalization or observation encounter from disease date through index.
- Plasma exchange, diffuse alveolar hemorrhage, chronic kidney disease, acute kidney injury, dialysis timing, kidney transplant, and other kidney manifestations.
- Recent and remote glucocorticoid measures, including 60-day average PEDD, recent and remote pulse glucocorticoids, and recent and remote high-dose oral glucocorticoids.
- Charlson Comorbidity Index (CCI), type 2 diabetes, hypertension, liver disease, serious infection, osteoporosis, and other inflammatory disease.
- Recent and remote rituximab, recent and remote cyclophosphamide, combined rituximab and cyclophosphamide use, and other immunosuppressive medication use.
- Outpatient visits and combined inpatient/emergency room visits.
- Disease event type (new vs relapsing disease).
- Healthcare access or engagement variables, including wellness visits, influenza vaccination, cholesterol testing, and cancer screening.

**Statistical Analysis**

Statistical analyses were conducted using the causalRisk R package version 0.41.2.7005.^1^ Propensity scores were estimated using logistic regression pooled across sequential trials. The propensity score was defined as the probability of initiating avacopan on the index date (A=1) versus not initiating avacopan (A=0), conditional on baseline covariates (W): $\hat{Pr}(A=1|W_{i}).$ The subscript *i* denotes values for individual *i*. Normalized standardized mortality ratio (SMR) weights were applied to reweight the SoC arm to the covariate distribution of avacopan initiators, targeting a treatment effect in the treated population, as follows:

For an individual in the **avacopan** arm, the weight is

$$\hat{z}_{i,a}= 1,$$

and for an individual in the **SoC** arm, the weight is

$$\hat{z}_{i,s}=\frac{\hat{Pr}(A=1|W_{i})}{(1- \hat{Pr}\left( A=1 | W_{i} \right))}.$$

These weights are then standardized within each treatment group to sum to the total sample size, *n*.

For the **avacopan arm,** these weights are

$$\hat{z}_{i,norm}=\hat{z}_{i,a}\frac{n}{\sum_{l=1}^{n_{a}} \hat{z}_{l,a}}$$

and for the **SoC arm**, they are

$$\hat{z}_{i,norm}=\hat{z}_{i,s}\frac{n}{\sum_{l=1}^{n_{s}} \hat{z}_{l,s}}.$$

Covariate balance was assessed using standardized mean differences before and after weighting, along with propensity score overlap and weight distributions.

Primary effectiveness analyses followed a modified intention-to-treat estimand. Under this estimand, SoC records were censored if avacopan was initiated within 30 days after index to account for short delays between treatment intent and the observed avacopan prescription. Inverse probability of censoring (IPC) weights were used for comparative effectiveness outcomes to account for potentially informative censoring due to disenrollment, death, administrative end of follow-up, and avacopan initiation within 30 days after index in the SoC arm. The time in days from the index date to the outcome was defined as *Y,* and the time in days from the index date to the time of censoring due to death, disenrollment, administrative end of follow-up, or initiation of avacopan within 30 days of index in the SoC arm was defined as *C.* *T=min(Y,C)* is the minimum of the time to the outcome and the time to censoring, and

$$\Delta= I\left( Y\leq C \right)$$

is an indicator that the outcome occurred before censoring or on the day of censoring.

The probability of remaining uncensored over time, conditional on treatment arm and measured baseline covariates that may affect censoring and the outcome, was estimated using main terms Cox proportional hazards models fit separately by treatment arm. For the smaller avacopan arm with the primary reason for potentially informative censoring being disenrollment, the censoring model included age, simplified insurance type, Charlson Comorbidity Index, total healthcare visits, recent pulse glucocorticoids, recent high-dose oral glucocorticoids, wellness visits, influenza vaccination, cholesterol testing, and cancer screening. For the SoC arm, the full propensity score covariate set was used for the censoring model to better address censoring related to post-index avacopan initiation. This choice was supported by negative control outcome results suggesting reduced bias when this censoring model was used as opposed to using the simplified censoring model for both arms, prior to analysis of study outcomes. We used these models to predict

$$\hat{Pr}\left( \Delta=1 | W_{i},A_{i},T_{i} \right).$$

This probability was used in the denominator of the IPC weights.

***Estimation of Hazard Ratios (HRs)***

Unweighted HRs: We estimated unweighted HRs from an unadjusted Cox proportional hazards model with treatment arm (*A*) as the exposure:

$$\lambda\left( t | A_{i} \right)=\lambda_{0}\left( t \right)\exp\left( A_{i}\beta\right)$$

where the HR is given by $exp(\beta)$ and $\lambda_{0}(t)$ is the baseline hazard.

Weighted HRs: We estimated SMR and IPC-weighted HRs from a marginal structural Cox proportional hazards model. Let

$$\hat{z}_{i,cens}= \frac{1}{\hat{Pr}(\Delta=1|W_{i},A_{i}, T_{i})}.$$

$exp(\beta)$ from this model is determined by maximizing the weighted partial likelihood^2^

$$L\left( \beta\right)= {\prod_{i=1}^{n} \left[ \frac{\exp\left( \beta A_{i} \right)}{\sum_{j\in R\left( t_{i} \right)} \hat{z}_{j,norm} \hat{z}_{j,cens}\exp\left( \beta A_{j} \right)} \right]}^{Y_{i}*\hat{z}_{i,norm}*\hat{z}_{i,cens}}$$

where $R(t)$ is the set of participants at risk at time *t.*

***Estimation of Risk Ratios (RRs):***

Unweighted RRs: For patients in a given treatment arm, the unadjusted cumulative risk of the outcome by time *t* was given by

$$\hat{Pr}\left( Y<t \right)= \frac{1}{n}\sum_{i=1}^{n} \frac{\Delta_{i}I(Y_{i}<t)}{\hat{Pr}(\Delta=1|T_{i})}$$

where *n* is the number of patients in the treatment arm for which the cumulative risk is being estimated.

The unadjusted relative risk by time *t* is given by

$$\hat{RR}\left( t \right)= \frac{\hat{Pr}(Y<t|A=1)}{\hat{Pr}(Y<t|A=0)}.$$

Weighted RRs:

The cumulative risk of the specified outcomes by 12 months under an intention-to-treat strategy of adding avacopan to SoC – adjusted for measured confounders using SMR weights and measured causes of informative censoring using IPC weights – is given by

$$\frac{1}{n}\sum_{i=1}^{n} \hat{z}_{i,norm}\frac{\Delta_{i}I\left( Y_{i}<t \right)I(A_{i}=1)}{\hat{Pr}\left( \Delta=1 | {W_{i}, A}_{i},T_{i} \right)}$$

where *n* is the total number of individuals in both arms.

The cumulative risk of the specified outcomes by 12-months under a modified intention-to-treat strategy of continuing SoC and not starting avacopan within 30 days after index – adjusted for measured confounders using SMR weights and measured causes of informative censoring using IPC weights – is given by

$$\frac{1}{n}\sum_{i=1}^{n} \hat{z}_{i,norm}\frac{\Delta_{i}I\left( Y_{i}<t \right)I(A_{i}=0)}{\hat{Pr}\left( \Delta=1 | {W_{i},A}_{i},T_{i} \right)}$$

where *n* is the total number of individuals in both arms. The relative risk is then the ratio of these cumulative risks by 12 months.

***Estimation of the Proportion of Individuals With PEDD ≤ 7.5 mg at 90, 120, 180, 270, and 365 days***

Unweighted Proportions:

We also estimated the proportion of individuals with 30-day average PEDD ≤ 7.5 mg at 90, 120, 180, 270, and 365 days by treatment arm. With $Y_{i}$ denoting the 30-day average PEDD at a specified timepoint the unweighted proportion with average PEDD ≤ 7.5mg for a specific treatment arm is given by

$$\hat{Pr}\left( Y\leq7.5 \right)= \frac{1}{n}\sum_{i=1}^{n} \frac{I(Y_{i}\leq7.5)I(C_{i}\geq t)}{\hat{Pr}(C\geq t)}$$

We also report the ratio of the arm-specific proportions.

Weighted Proportions:

We estimated the weighted proportion with 30-day average PEDD ≤ 7.5mg at 90, 120, 180, 270, and 365 days. For this analysis, we used the same censoring models described above to predict the probability of remaining uncensored through the timepoint of interest: $\hat{Pr}(C\geq t|W_{i}, A_{i})$. The estimates of the proportions with 30-day average PEDD ≤ 7.5mg at the specified timepoints for the avacopan and SoC arms were given by:

$$\frac{1}{n}\sum_{i=1}^{n} \hat{z}_{i,norm}\frac{I\left( Y_{i}\leq7.5 \right)I(A_{i}=1, C_{i}\geq t)}{\hat{Pr}\left( C\geq t | {W_{i}, A}_{i} \right)}$$

and

$$\frac{1}{n}\sum_{i=1}^{n} \hat{z}_{i,norm}\frac{I\left( Y_{i}\leq7.5 \right)I(A_{i}=0, C_{i}\geq t)}{\hat{Pr}\left( C\geq t | {W_{i}, A}_{i} \right)}$$

The relative risk is then the ratio of these proportions.

***Estimation of the difference in mean 30-day average PEDD at 0, 30, 60, 90, 120, 150, 180, 210, 240, 270, 300, 330, and 360 days***

Unweighted Differences:

We estimated the 30-day average PEDD at 30, 60, 90, 120, 150, 180, 210, 240, 270, 300, 330, and 360 days by treatment arm. With $Y_{i}$ denoting the 30-day average PEDD at a specified timepoint, the mean 30-day average PEDD is given by

$$\hat{E}[Y_{i}]= \frac{1}{n}\sum_{i=1}^{n} \frac{Y_{i}I(C_{i}\geq t)}{\hat{Pr}(C\geq t)}$$

Weighted Differences:

We estimated the SMR- and IPC-weighted mean 30-day average PEDD at 0, 30, 60, 90, 120, 150, 180, 210, 240, 270, 300, 330, and 360 days. Using the same censoring models described above, we predicted the probability of remaining uncensored through the timepoint of interest: $\hat{Pr}(\Delta_{t}=1|W_{i}, A_{i})$, where $\Delta_{t}=I(C\geq t)$. With $Y_{i}$ representing the 30-day average PEDD at a specified timepoint, the estimates of the mean 30-day average PEDD at the specified timepoints for the avacopan and SoC arms were given by:

$$\frac{1}{n_{1}^{*}}\sum_{i=1}^{n} \hat{z}_{i,norm}\frac{Y_{i}*I(A_{i}=1, C_{i}\geq t)}{\hat{Pr}\left( C\geq t | {W_{i}, A}_{i} \right)}$$

and

$$\frac{1}{n_{0}^{*}}\sum_{i=1}^{n} \hat{z}_{i,norm}\frac{Y_{i}*I(A_{i}=0, C_{i}\geq t)}{\hat{Pr}\left( C\geq t | {W_{i}, A}_{i} \right)}$$

where $n_{a}^{*}$ is the sum of the weights given by

$$n_{a}^{*}= \sum_{i=1}^{n} \hat{z}_{i,norm}\frac{I(A_{i}=a, C_{i}\geq t)}{\hat{Pr}\left( C\geq t | {W_{i}, A}_{i} \right)}$$

The difference in means is then the difference between these averages.

***Estimation of the difference in mean cumulative glucocorticoid exposure at 30, 60, 90, 120, 150, 180, 210, 240, 270, 300, 330, and 360 days.***

Unweighted Difference in Means:

We estimated the mean cumulative glucocorticoid exposure at 30, 60, 90, 120, 150, 180, 210, 240, 270, 300, 330, and 360 days by treatment arm. With $Y_{i,k}$ denoting the PEDD on post-index day *k*, t denoting the timepoint of interest, and $\Delta_{i,k}=I\left( C_{i}\geq k \right),$the mean cumulative glucocorticoid exposure is given by

$$\frac{1}{n}\sum_{i=1}^{n} \sum_{k=1}^{t} \frac{{\Delta_{i,k}Y}_{i,k}}{\hat{Pr}(\Delta_{k}=1)}$$

Weighted Difference in Means:

We estimated the SMR- and IPC-weighted mean cumulative glucocorticoid exposure at 30, 60, 90, 120, 150, 180, 210, 240, 270, 300, 330, and 360 days. Using the same censoring models described above, we predicted the probability of remaining uncensored through the timepoint of interest: $\hat{Pr}(\Delta_{t}=1|W_{i}, A_{i})$, where $\Delta_{t}=I(C\geq t)$. With $Y_{i,k}$ representing the PEDD on post-index day *k*, the estimates of the mean cumulative glucocorticoid exposure at the specified timepoints for the avacopan and SoC arms were given by

$$\frac{1}{n}\sum_{i=1}^{n} \sum_{k=1}^{t} \hat{z}_{i,norm}\frac{Y_{i,k}*I(A_{i}=1, \Delta_{i,k}=1)}{\hat{Pr}\left( \Delta_{k}=1 | {W_{i}, A}_{i} \right)}$$

and

$$\frac{1}{n}\sum_{i=1}^{n} \sum_{k=1}^{t} \hat{z}_{i,norm}\frac{Y_{i,k}*I(A_{i}=0, \Delta_{i,k}=1)}{\hat{Pr}\left( \Delta_{k}=1 | {W_{i}, A}_{i} \right)}$$

The difference in means is then the difference between these averages.

For hepatic safety endpoints in the effectiveness cohort, 12-month cumulative risk was estimated separately by treatment arm using baseline propensity score SMR weights without IPC weights. In addition, a separate descriptive avacopan safety cohort included adults with any recorded avacopan prescription during observed enrollment within the study period, regardless of whether they met eligibility criteria for the effectiveness analysis. For this cohort, the number and proportion of patients with a serious hepatic event within 1 year after first recorded avacopan prescription were summarized descriptively.

95% confidence intervals were estimated based on the 2.5^th^ and 97.5^th^ percentiles of an individual-level Bayesian bootstrap distribution of the target parameter with Dirichlet weights. Bootstrap resampling was performed at the patient level to account for intrapatient correlation from patients contributing more than one eligible index date. The primary and secondary endpoint analyses used 250 bootstrap iterations; negative control analyses used 100 iterations.

**Negative Control Outcomes and Clean-room Process**

Negative control outcomes were used to assess potential residual bias after propensity score and censoring adjustment. The negative control outcomes relevant to the manuscript were preventive cancer screening and wellness visits. Weighted and unweighted 12-month cumulative risk curves, risk ratios, and risk differences were evaluated before primary and secondary outcome analyses were conducted.

The analysis was conducted using a staged clean-room process.^3^ Initial stages included cohort feasibility assessment, unweighted and weighted baseline characteristics, propensity score diagnostics, and negative control outcome analyses. Predefined checkpoints were used to determine whether inclusion criteria, propensity score models, censoring models, or the SoC indexing approach required revision before comparative effectiveness outcomes were analyzed. Treatment effect estimates were not reviewed until after the final analysis specifications were locked. The process also included restricted access to analytic folders, masking of treatment-group information where appropriate, double programming or quality control of key analyses, and documentation of decisions in an audit trail.

**Supplementary Coding Summary**

The following summary lists key diagnosis, medication, procedure, and outcome coding categories used in the analyses. Specific ICD-10-CM codes are shown where the code list is brief; broader medication and procedure definitions were implemented using relevant diagnosis, procedure, HCPCS/CPT, and/or NDC codes.

| Concept | Claims codes or code category |
| --- | --- |
| GPA | ICD-10-CM: M31.3, M31.30, M31.31 |
| MPA | ICD-10-CM: M31.7 |
| AAV | ICD-10-CM: I77.82 |
| EGPA | ICD-10-CM: M30.1 |
| Avacopan | NDC codes for avacopan |
| Rituximab | NDC, HCPCS/CPT, and procedure codes for rituximab |
| Cyclophosphamide | NDC, HCPCS/CPT, and procedure codes for cyclophosphamide |
| Strict hepatic event screen, criterion 1 | ICD-10-CM diagnosis codes for acute/subacute hepatic failure, central hemorrhagic necrosis of liver, or toxic liver disease |
| Strict hepatic event screen, criterion 2 | ICD-10-CM diagnosis codes indicative of severe acute liver injury |

Code lists were implemented programmatically as ATLAS concept sets for the analysis. Because numeric ATLAS concept set identifiers are implementation-specific, this supplement reports the clinical concept, coding system, and selected code values where applicable rather than internal concept set identifiers.

**Prednisone-equivalent Conversion Factors**

| Glucocorticoid | Equivalent dose (mg) | Conversion factor to prednisone |
| --- | --- | --- |
| Prednisone | 5 | 1.00 |
| Prednisolone | 5 | 1.00 |
| Methylprednisolone | 4 | 1.25 |
| Dexamethasone | 0.67 | 7.47 |

**eTable 1. Modified ITT Estimands for Comparative Effectiveness and Safety**

| **Target Trial Protocol Component** | **Target Trial Specification** | **Target Trial Emulation** |
| --- | --- | --- |
| **Eligibility Criteria** | - New or relapsing GPA/MPA within the past 125 days - Age ≥18 years - Recent rituximab (within 90 days) and/or cyclophosphamide (within 45 days) - No prior avacopan use - No prior diagnosis of EGPA - No prior use of mepolizumab or benralizumab   The target population is the population represented by the avacopan-treated patients (treatment effect in the treated) | - Patients meeting the criteria for new or relapsing GPA/MPA within 125 days prior to the index date using the specified claims-based algorithms - Age ≥18 years at index - ≥1 claim for rituximab within 90 days and/or cyclophosphamide within 45 days prior to or on the index date - No filled prescriptions for avacopan prior to the index date, using all lookback - No diagnosis of EGPA in the lookback period - No filled prescriptions for mepolizumab or benralizumab in the lookback period - 1 year of continuous medical and pharmacy enrollment prior to the index date with ≤30-day gap - At least 1 day of follow-up   The target population is the population represented by the avacopan-treated patients (treatment effect in the treated) |
| **Treatment Strategies** | Among the eligible patients who are receiving SoC:  1. Initiate avacopan at time zero  2. Do not initiate avacopan at time zero or for 30 days after time zero | Among the eligible patients who meet the claims-based criteria for receiving SoC:  1. Fill one prescription for avacopan on the index date  2. Do not fill a prescription for avacopan on the index date or for 30 days after the index date |
| **Assignment Procedures** | Randomization | Pseudo-randomization mimicked using SMR weights to balance measured confounders at baseline |
| **Follow-up Period** | Randomization to loss to follow-up, the outcome (including death), or 12 months | The day after the index date to disenrollment, end of administrative follow-up, the outcome (including death), or 12 months  Index date: Sequential nested trials will be emulated every 14 days starting from the disease date for new or relapsing GPA/MPA. Within each trial, index dates will be defined as:   - Avacopan arm: Prescription fill date for avacopan - SoC arm: Two methods of defining the SoC index date will be considered. The final method will be chosen based on unweighted and weighted balance of baseline covariates, propensity scores, and, potentially, NCO results prior to running any outcomes analyses^a^   –Method 1: The first date of either a claim for rituximab, a claim for cyclophosphamide, or an encounter with a nephrologist or rheumatologist in the interval  –Method 2: The first date of either a claim for rituximab or a claim for cyclophosphamide in the interval |
| **Outcomes** | Primary endpoints:   - Time to first relapse - 30-day average PEDD ≤7.5 mg at 90, 120, 180, 270, and 365 days   Secondary endpoints:   - Relapse within 12 months - Hepatotoxicity and drug-induced liver injury   Post-hoc endpoints:   - 30-day average PEDD (and change from baseline 30-day average PEDD) by 30, 60, 90, 120, 150, 180, 210, 240, 270, 300, 330, 360 days - Cumulative glucocorticoid exposure by 30, 60, 90, 120, 150, 180, 210, 240, 270, 300, 330, 360 days | Same as in target trial, as captured in claims data |
| **Causal Contrasts** | Effects will be summarized as cumulative risk by arm, proportion by arm, risk ratios, risk differences, or HRs, as appropriate for the outcome. We estimate treatment effects in treated patients (avacopan initiators) | Effects will be summarized as cumulative risk by arm, proportion by arm, risk ratios, risk differences, or HRs, as appropriate for the outcome. We estimate treatment effects in treated patients (avacopan initiators) |

^a^Method 2 was chosen based on propensity score diagnostics, unweighted and weighted baseline characteristics, and NCO analyses.

EGPA, eosinophilic granulomatosis with polyangiitis; ESKD, end-stage kidney disease; GPA, granulomatosis with polyangiitis; HR, hazard ratio; ITT, intent-to-treat; MPA, microscopic polyangiitis; NCO, negative control outcomes; PEDD, prednisone-equivalent daily dose; SMR, standardized mortality ratio; SoC, standard of care.

**eFigure 1.** **Study Design**

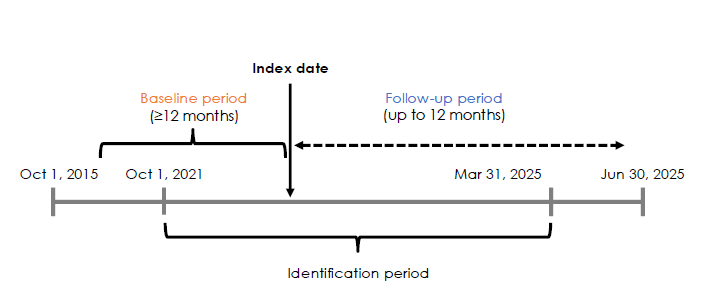

**eFigure 2.** **Patient Attrition**

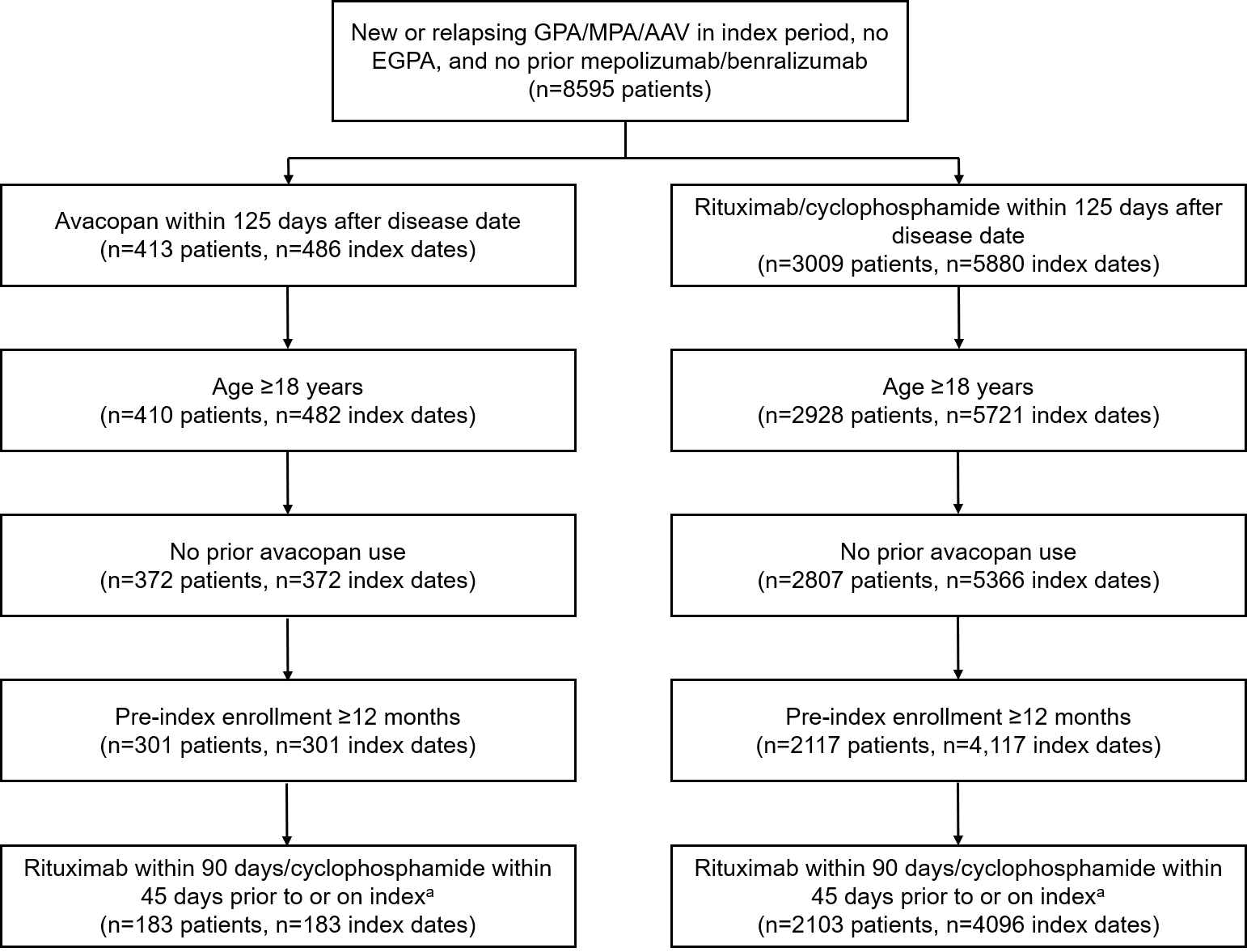

^a^Excluded unknown gender or insurance; no death date on or before index date and no disenrollment on index date.

AAV, antineutrophil cytoplasmic antibody–associated vasculitis; EGPA, eosinophilic granulomatosis with polyangiitis; GPA, granulomatosis with polyangiitis; MPA, microscopic polyangiitis.

**eFigure 3.** **Standardized Mean Differences of Key Baseline Characteristics Among the Avacopan Plus SoC and SoC Arms Before and After Propensity Score Weighting**

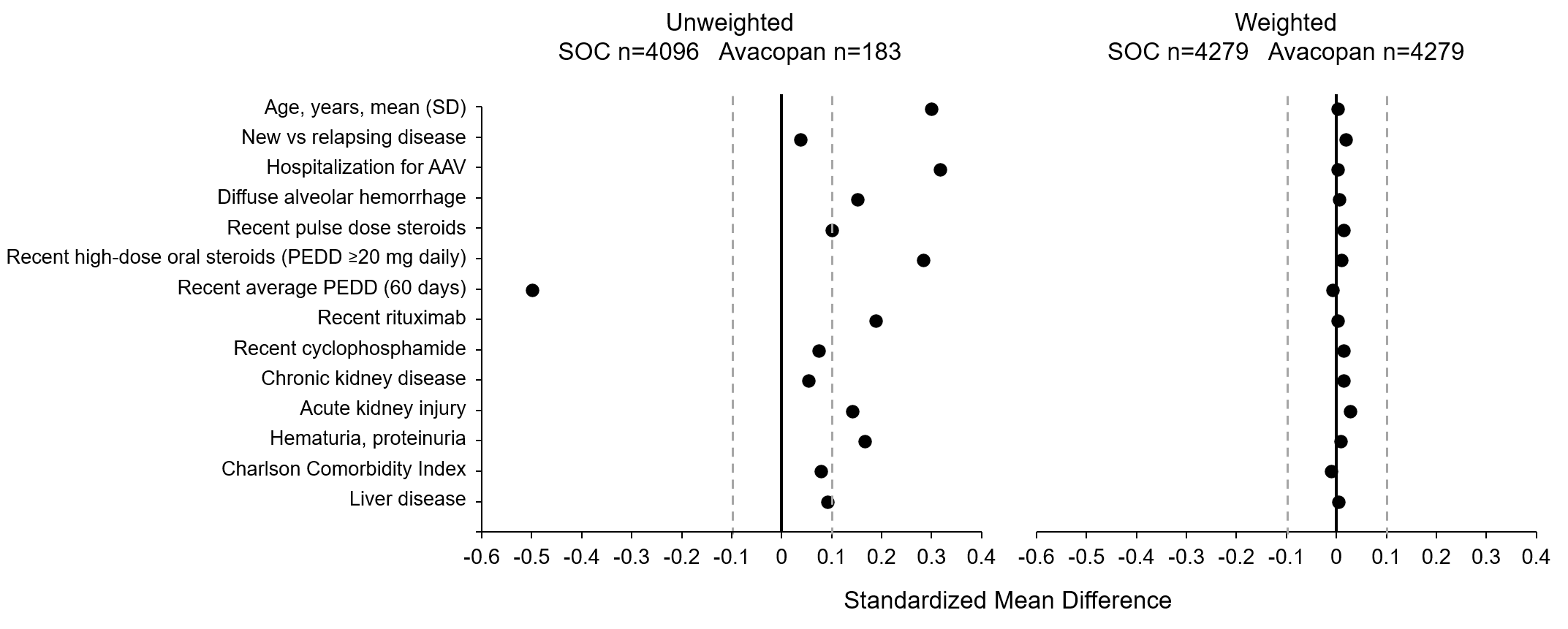

Weighted n is equal to the sum of the normalized standard mortality ratio weights, which are normalized to sum to the total sample size.

AAV, antineutrophil cytoplasmic antibody–associated vasculitis; PEDD, prednisone-equivalent daily dose; SD, standard deviation; SoC, standard of care.
